# An Interactive Tool to Forecast US Hospital Needs in the Coronavirus 2019 Pandemic

**DOI:** 10.1101/2020.04.20.20073031

**Authors:** Kenneth J. Locey, Thomas A. Webb, Jawad Khan, Anuja K. Antony, Bala Hota

## Abstract

Hospital enterprises have been continually faced with anticipating the spread of COVID- 19 and the effects it is having on visits, admissions, bed needs, and crucial supplies. While many studies have focused on understanding the basic epidemiology of the disease, few open source tools have been made available to aid hospitals in their planning. We developed a web-based application (available at: http://covid19forecast.rush.edu/) for US states and territories that allows users to choose from a suite of models already employed in characterizing the spread of COVID-19. Users can obtain forecasts for hospital visits and admissions as well as anticipated needs for ICU and non-ICU beds, ventilators, and personal protective equipment supplies. Users can also customize a large set of inputs, view the variability in forecasts over time, and download forecast data. We describe our web application and its models in detail and provide recommendations and caveats for its use. Our application is primarily designed for hospital leaders, healthcare workers, and government official who may lack specialized knowledge in epidemiology and modeling. However, specialists can also use our open source code as a platform for modification and deeper study. As the dynamics of COVID-19 change, our application will also change to meet emerging needs of the healthcare community.

## INTRODUCTION

Coronavirus disease 2019 (COVID-19) was declared a global pandemic by the World Health Organization on March 11^th^ 2020. By then, confirmed COVID-19 cases were reported among 109 countries and exceeded 128,000 worldwide (data source: Johns Hopkins University Center for Systems Science and Engineering). That number increased more than 10-fold in less than one month. As COVID-19 spreads within and among nations, healthcare enterprises grapple with the challenges of preparing for the growing number of COVID-19 patients and with appropriating the resources needed to treat patients while protecting healthcare professionals. In the United States (US), policy makers and hospital leaders prognosticate on how to best allocate resources in the face of an anticipated surge in demand that may last for several months to come (Bukhari and Jameel, 2020). However, even as COVID-19 threatens to overwhelm healthcare systems, the predictive analytics tools that would otherwise allow hospitals to make informed decisions are lagging behind the increasing number of studies aimed at characterizing the basic epidemiology of COVID-19.

To meet the pressing needs of hospital enterprises we developed an interactive, open-source web-based application to provide state- and hospital-specific forecasts of COVID-19 patients and related supplies. Our application is available on the http://covid19forecast.rush.edu/ website and allows users to employ a suite of models already used in the prediction of COVID-19 cases. It then couples these models to granular customizable inputs to produce hospital-level forecasts for COVID-19 visits and admits, ICU and non-ICU beds, ventilators, and various personal protective equipment (PPE) supply needs. Our efforts are aimed at addressing immediate and anticipated healthcare demands, and to allow informed decision-making by government officials and healthcare professionals who may lack specialized expertise in epidemiology, modeling, and data science. By making our aggregated data and source code freely available, and by offering additional source code outside the application itself, epidemiologists, modelers, and data scientists may also find our application useful as is, or as a modifiable resource for deeper analysis.

In this paper, we describe our application in detail, focusing on the data and models it uses, the inputs it allows users to enter, and the graphs, tables, and downloadable data it provides. We also provide guidance on the use of this application, the interpretation of its outputs, and the caveats of our approach. Finally, we discuss the value of our application to meeting the needs of the healthcare community, its potential as a tool for generating novel insights, and modifications to come. In addition to our aim of empowering administrators, physicians, and governmental agencies to make informed decisions, we sought to enable other predictive healthcare analytics teams and researchers. Specifically, the modification and deployment of our application’s source code requires a minimal set of widely popular open source software (e.g., python language, Jupyter notebook) and little-to-no experience in web development.

## FUNCTIONALITY AND USE

### Overview

Our open-source application allows users 1) to aggregate data from a popular open-source repository of COVID-19 data, 2) to track and forecast the progression of COVID-19 cases across US states using a suite of well-known models, 3) customize a large set of input parameters to provide state- and hospital-specific forecasts for numbers of hospital visits, admitted patients, ICU needs, and personal protective equipment (PPE) supply needs. The application also allows users to adjust the length of forecasts, to adjust expected time lags in patient visits, to adjust the average length of stay (LOS) for ICU and non-ICU patients, to examine forecasts from previous days, and to download forecast data for deeper analysis.

### Data

Our application accesses COVID-19 data from the Johns Hopkins University Center for Systems Science and Engineering (JHU CSSE) (2). Specifically, our application downloads, aggregates, and stores daily reports from the JHU CSSE public GitHub repository (https://github.com/CSSEGISandData/COVID-19). These daily reports contain cumulative numbers of confirmed cases, and cumulative numbers of reported deaths and recoveries for counties, states, provinces, and nations reported since January 22^nd^, 2020. For select models, our application uses population sizes for US states and territories based on data from the US Census Bureau (2010 – 2019). Our application also uses dates of COVID-19 arrival in US states and territories based on data available from state and territory governmental agencies (e.g., Departments of Health).

### Modeling COVID-19 cases

Our application begins by modeling the cumulative COVID-19 cases for a selected state, and allows users to choose from four simplistic models of growth that have been used in characterizing the spread of COVID-19 as well as one popular epidemiological model that has been frequently used in the study of COVID-19 (Fig 1). Altogether, the following suite of models allows users the ability to compare and contrast forecasts resulting from different forms of growth and varying complexity of disease dynamics and social response (e.g., testing lags, social distancing).

**Figure 1.**
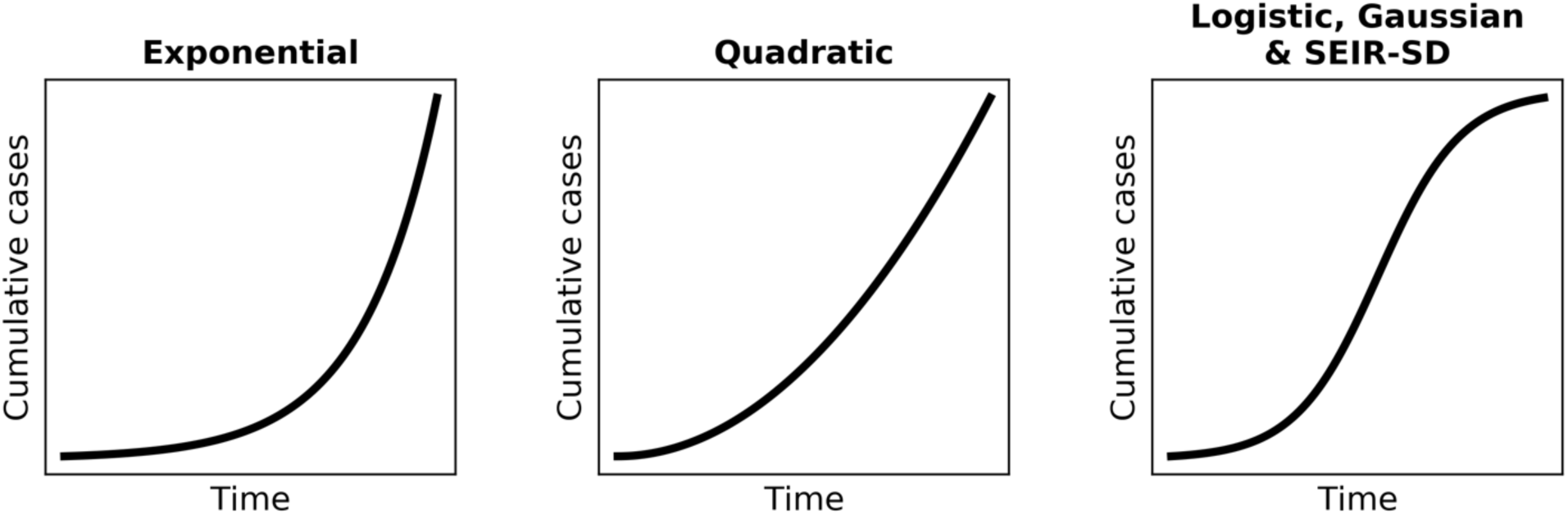
Our application allows users to choose from five models that have previously been employed in the study of COVID-19, the general cumulative forms of which are depicted below. While the exponential and quadratic (2^nd^ degree polynomial) models only allow for continued increase in cumulative cases, the logistic, Gaussian, and SEIR-SD models allow cumulative cases to saturate.

#### Exponential growth

Initial stages of growth often appear limited only by the inherent growth rate (*r*) of the population or disease. In this way, exponential growth proceeds multiplicatively according to a simple functional form, *N_t_* = *N_0_e^rt^*, where *N_0_* is the initial infected population size, *t* is the amount of passed, and *N_t_* is the infected population size at *t*. The exponential model has been widely used to characterize the spread of COVID-19 the during initial weeks of infection (e.g., Graselli, Presenti, and Cecconi 2020, Lui *et al*. 2020, Remuzzi and Remuzzi, 2020; Fig 2). Because it assumes that *r* is constant, the exponential model has a simple log-linear transformation, log(*N_t_*) = log(*N_0_*) + *t* • *r*, that allows log-transformed numbers of cases to be regressed on *t* (Sit, Poulin-Costello, and Bergerud 1994). Our application uses this exponential regression to obtain predictions for the expected cumulative number of confirmed COVID-19 cases (*N*). This model has explained upwards of 99% of variation in the initial days or weeks of COVID-19 spread within states; however, it quickly begins to fail because it only allows for continued rapid growth (Fig 2).

**Figure 2.**
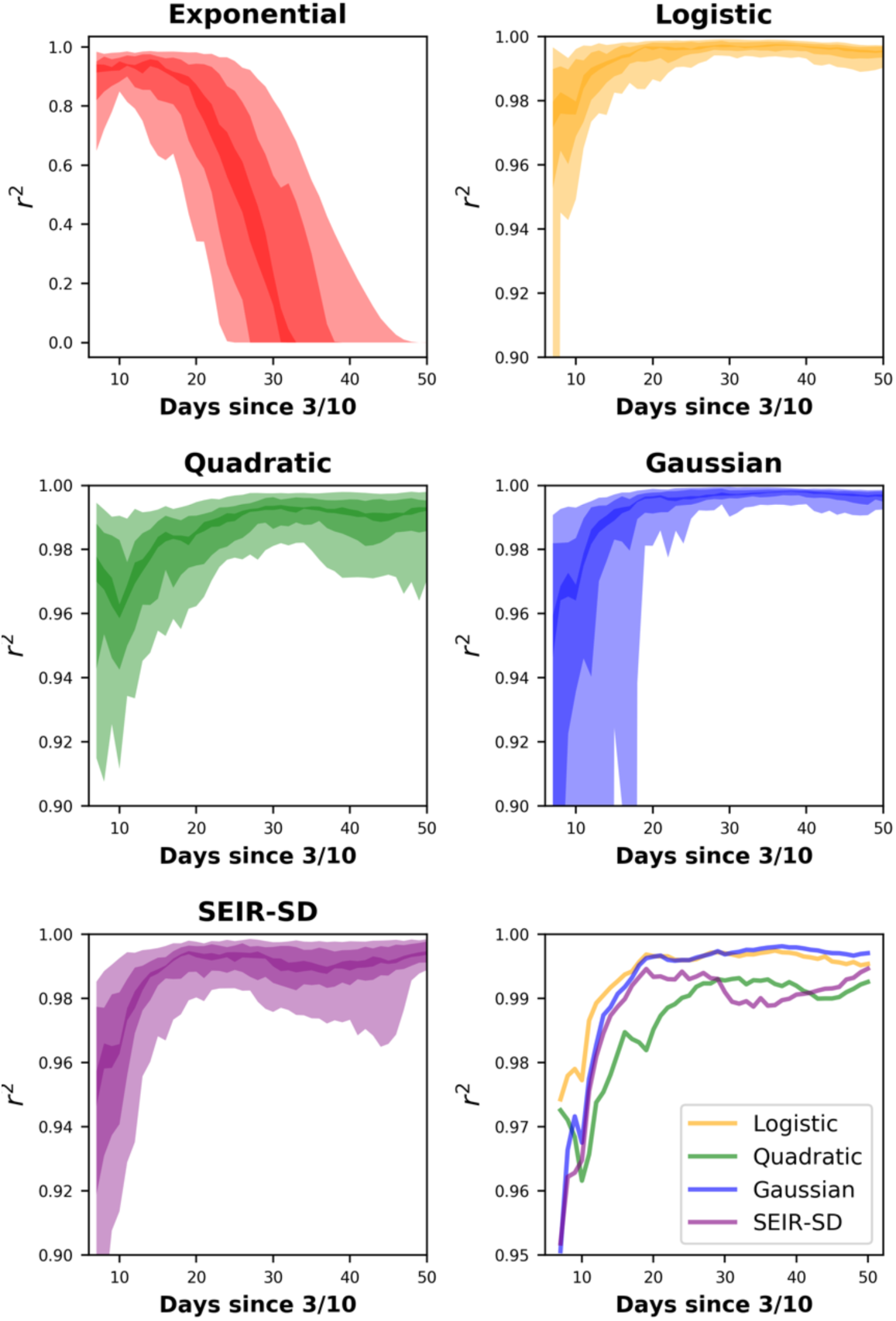
Confidence interval (CI) hulls for the performance of models across US states and territories from March 16th to April 15th. The Johns Hopkins University Center for Systems Science and Engineering (JH CSSE) data shows most states beginning to report COVID-19 cases by Marth 10th and we allowed the models a week of data before fitting them. The *x*-axis is in days since the first reported case, occurring on or after March 10th. Lightly colored hulls are 90% CI, hulls of intermediate darkness are 50%, and the darkest hulls are 10% CI. Performance is measured via a modified *r*^2^ of observed versus predicted values where the *y*-intercept is forced through 0 (*sensu* Locey and White 2013). The y-axes are scaled to show the greatest resolution for each model and are not scaled similarly across all models. Lower right: Mean *r*^2^ values among states for all models except the exponential. This figure reveals the accuracy of models fitted to observed data, but do not pertain to the accuracy of future forecasts.

#### Quadratic growth

Initial stages of growth may be more rapid than that expected from the exponential model while the latter monotonic increase in *N* can proceed less rapidly than predicted by the exponential. In these cases, growth may be quadratic, i.e., characterized by a constant change in growth rate. Early COVID-19 studies have implicated quadratic growth in spread of COVID-19 (e.g., Brandenburg 2020; Fang, Ne, and Penny 2020) and the quadratic model, to date, has continued to perform well (Fig 2). The quadratic function, *f*(*x*) = *x*^2^ + *x* + *c*, is a 2^nd^ order polynomial that can be applied to population growth as *N_t_* = *β*_1_*t*^2^ + *β*_2_*t* + *N_0_*. Our application uses numerical optimization of the fitted parameters, *β*_1_ and *β*_2_, to find the best fit quadratic function for a given time series and hence, to predict values for (*N*). This model has, thus far, improved as COVID-19 spreads and has explained upwards of 99% of variation in COVID-19 cases among states (Fig 2). However, like the exponential model, the quadratic model only allows for continued growth, i.e., no saturation. Consequently, the quadratic model must eventually fail as COVID-19 cases saturate.

#### Logistic growth

Exponential growth within a population cannot continue *ad infinitum*. Instead, growth must slow as an upper limit is approached or as natural limitations to disease spread (e.g., immunity, contact among hosts) are encountered. The logistic model captures this slowing and eventual saturation, resulting in a sigmoidal or s-shaped growth curve (Maynard-Smith 1978, Bacaër 2011, Fig 1). In addition to exponential and quadratic growth, early COVID-19 studies have implicated logistic growth in the spread of the disease (Roosa *et al*. 2020, Wu *et al*. 2020). Like the quadratic model, the logistic model has also continued to perform well as states have progressed in COVID-19 infection (Fig 2). The logistic model takes a relatively simple functional form, 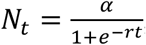, where *α* is the upper limit of *N* and *r* is the intrinsic rate of increase. Our application uses numerical optimization of *α* and *r* to find the best fit logistic function and hence, predicted values for *N*. This model has, to date, improved as COVID-19 spreads and has explained upwards of 99% of variation in COVID-19 cases among states (Fig 2).

#### Gaussian growth

The Gaussian (i.e., normal) distribution can provide a relatively simple and close approximation to complex epidemiological models (Buckingham-Jeffery *et al*. 2018):

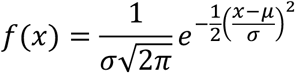

This symmetrical curve has two parameters, mean = *μ*, standard deviation = *σ*, and belongs to the family of exponential distributions (Fig 1). When used to model spread of disease, Gaussian curves are symmetrical around a climax day with the change in the rate of growth determining the standard deviation about the curve. Gaussian models have previously been successful in approximating the spread of COVID-19 in Germany (Schlickeiser and Schlickeiser 2020). Our application uses numerical optimization of *μ* and *σ* and the cumulative distribution function of the Gaussian model to find the best fit cumulative Gaussian function and hence, predicted values for *N*. This model has, thus far, explained >99% of variation in COVID-19 cases among states and continues to improve as COVID-19 spreads (Fig 2).

#### SEIR-SD

To date, COVID-19 studies have used a variety of epidemiological models to characterize the spread of the disease within populations. The modeling in several of these studies has been based on refinements to the classic SEIR model of Hethcote and Tudor (1980) (see Boldog *et al*. 2020, Peng *et al*. 2020, Lui *et al*. 2020, Wang *et al*. 2020). In this model, a contagious disease drives changes in the fraction of susceptible persons (*S*), the fraction of persons exposed but not yet exhibiting infection (*E*), the fraction of infectious persons (*I*), and the fraction of persons recovered (*R*), where *S* + *E* + *I* + *R* = 1. These SEIR subpopulations are modeled as compartments in the following set of ordinary differential equations:

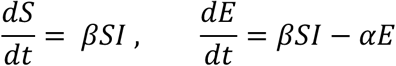

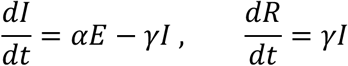

In these equations, *α* is the inverse of the incubation period, and *γ* is the inverse of the average infectious period, and *β* is the average number of contacts of infected persons with susceptible persons per unit time. Our application imputes the initial value of *β* from a well-known simplifying relationship between *γ* and the basic reproductive number (*R_0_*), i.e., *β = γ R_0_* (Ridenhour *et al*. 2018, Rocklöv *et al*. 2020, Rãdulescu and Cavanagh 2020, Lin et al. 2020).

We allowed *β* to decrease in proportion to *I*. We assumed that people will, on average, reduce their contact with others when the populace is aware that an increasing percent of their population is infected. This approach allows an inherent degree of social distancing to emerge as a frequency-dependent phenomenon. We also simulated an explicit effect of social distancing (*λ*) to capture the overall strength of response to public health policies. These effects were included as time-iterative modifications to *β*:

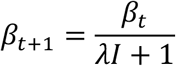

This function allows *β* to remain unchanged when either *I* or *λ* equal 0. When *λ* equals 1, the daily change in *β* is governed by the implicit frequency-dependent effect of *I*. Importantly, simple algebraic rearrangement shows that the product of social distancing (*λ*) and the fraction infected (*I*) determines the percent daily change in contact rate (*β*):

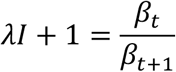

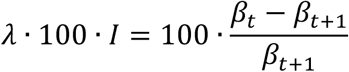

As a result, *λ* determines the daily proportional change in the contact rate per infected fraction of the total population:

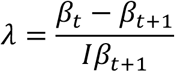

We also modified the classic SEIR model to account for initial time lags in COVID-19 testing. Specifically, and particularly in the US, widespread testing for COVID-19 may have artificially dampened the apparent number of positive cases within the first month of the first reported infection. We accounted for this effect by modifying the apparent size of *I* while allowing the actual size of *I* to grow according to the SEIR-SD dynamic. This time-iterative modification took the following logistic functional form:

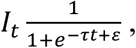

where *τ* and *ε* are fitted parameters. This equation models testing as low-to-nonexistent during the initial weeks of outbreak, and then accelerates afterwards.

To date, the SEIR-SD model has generally performed as well as or better than the exponential, quadratic, logistic, and gaussian models (Fig 2). Our application performs a pseudo-optimization on the SEIR-SD model parameters and a likely date of initial infection, as opposed to using the first reported occurrence. Our implementation of the SEIR-SD model is based on an unbiased search of multivariate parameter space within ranges of parameter values derived from population sizes for US states and territories and the increasing corpus of COVID-19 literature (Table 1). Our application performs 50,000 iterations and chooses the set of parameters that maximize the explained variation in observed data. This implementation avoids the computational challenges of applying numerical optimizers to complex simulation models and avoids the problems that these optimizers can have in becoming trapped in local minima.

**Table 1.** Biological parameters for the SEIR-SD model. Our application attempts to optimize the following SEIR-SD parameters within ranges of reported values for the average incubation period, the average infectious period, and the basic reproductive number.

| Parameter | Description | Reported Range | References |
| --- | --- | --- | --- |
| Avg. Incubation period (latent period) | Days during which an infected individual cannot infect others. | 5 – 6 days | Anderson *et al.* 2020<br>Pederson and Meneghini 2020<br>Wang *et al.* 2020<br>Kucharski *et al.* 2020<br>Cao *et al.* 2020 |
| Avg. Infectious period | Days over which an infected person remains infective. | 1.6 – 13 days | You *et al.* 2020<br>Maier and Brockmann 2020<br>Wang *et al.* 2020<br>Read *et al.* 2020 |
| Basic Reproductive Number ($R_0$) | Average number of secondary infections generated by a primary infection at the onset of outbreak | 2.1 – 6.5 | You *et al.* 2020<br>Pederson and Meneghini 2020<br>Maier and Brockmann 2020<br>Wang *et al.* 2020<br>Lui *et al.* 2020 |

### Forecasting COVID-19 cases

Our application allows users to select one of the above-mentioned models and a location from a list of US states and territories (Fig 3). It then plots the reported number of cumulative COVID-19 cases along with model predictions (up to present day) and forecasts (up to 60 days ahead) (Fig 3). Users can also view how the predictions and forecasts have changed over the last 10 days (Fig 3).

**Figure 3.**
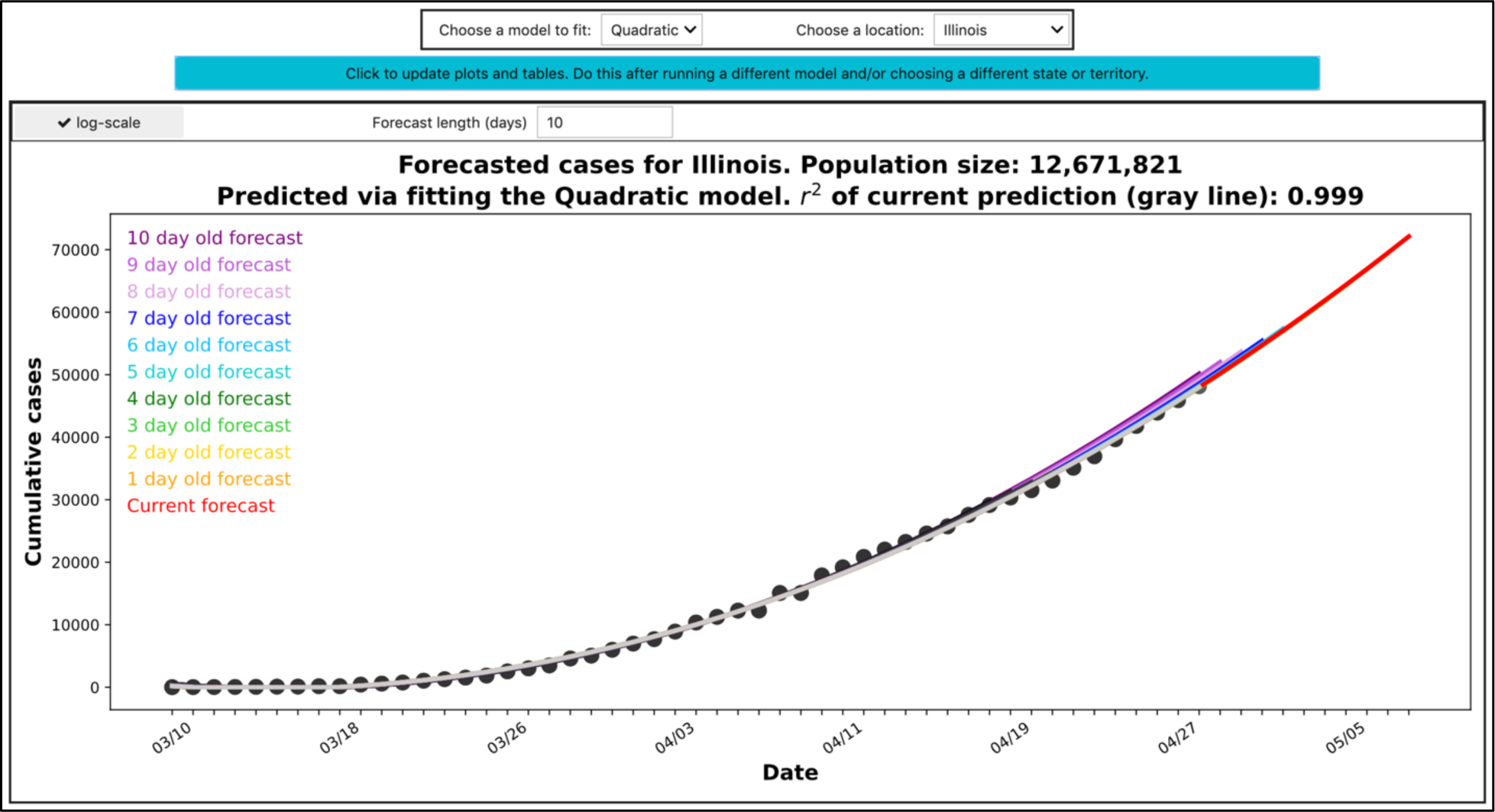
The top pane of the application features the ability to choose a location and one of four models for fitting data (black dots) and for making forecasts. The user then clicks a teal button to update the application’s plots of fitted predictions (gray-scale lines) and forecasts (colored lines). The application allows the user to extend the forecast window to 60 days and view logarithmically transformed data. The current image is the result of fitting the quadratic model to Illinois COVID-19 data and includes a forecast window of 10 days. Forecasts up to 10 days old are displayed to aid users in examining forecast variability. Values for coefficients of determination (*r*^2^) pertain to observed versus predicted values, where the *y*-intercept is forced through 0 (*sensu* Locey and White 2013).

### Forecasting hospital visits and admits via time lags

Our application allows users to enter the expected percent of state-wide COVID-19 cases that visit their hospital as well as the expected percent of those admitted. Going further, we accounted for the tendency of infected persons to not seek immediate medical attention. We modeled these time lags as Poisson distributed random variables, whereby portions of newly infected patients may wait 1, 2, 3, …, days to visit the hospital. The Poisson distribution has ideal properties for modeling this scenario. Specifically, it is a discrete probability distribution (*x*-values correspond to days) with a simplistic formulation (e.g., the mean equals the variance), where the only parameter needed to obtain the probability mass function (i.e., mean), directly corresponds to the expected average time lag. Users can view the change in the forecasted patient census by adjusting the expected time lag (Fig 4).

**Figure 4.**
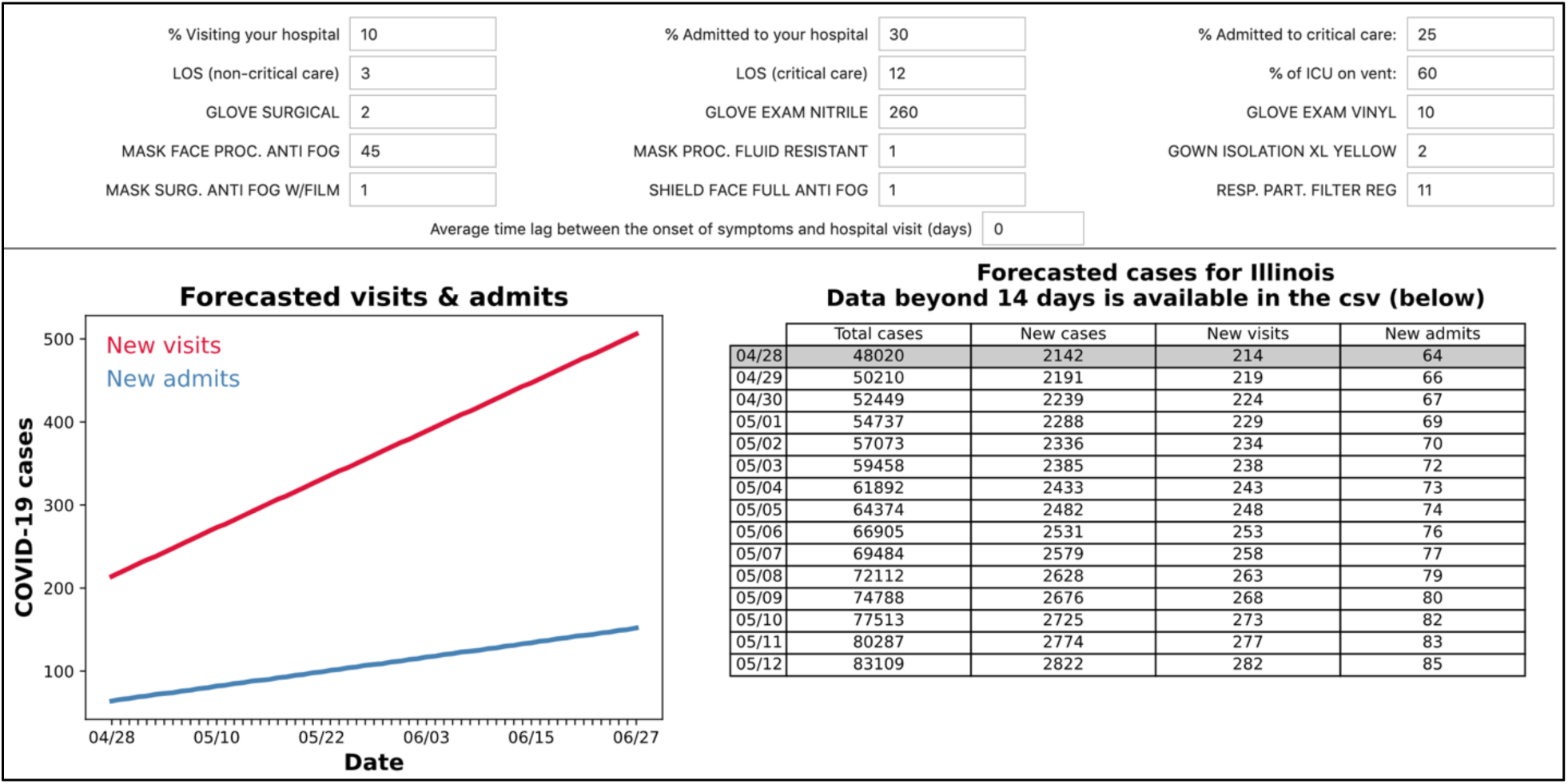
The second pane of our application allows users to set values for the percent of COVID-19 patients in a given state visiting their hospital, the percent admitted, and the percent admitted to critical care. Users can also enter the expected length of stay (LOS) for critical patients (ICU), non-critical care patient, the percent of ICU patients expected to be on ventilators, values for various PPE supply needs, and an expected time lag between when patients first experience symptom and when they visit the hospital. The application then plots the forecasted numbers of new visits and new admits, and then tabulates the forecasted total cases, new cases, new visits, and new admits. See figures 4 and 5 for plots and tables of patient census and PPE needs. The application informs the user that forecasted data beyond 14 days are available via csv downloads.

### Forecasting hospital bed and ICU needs via daily carry-over

Our application allows users to forecast the number of hospital beds, ICU beds, and ventilators needed. The user begins by entering expected values for the percent of COVID-19 patients admitted to critical care, the expected average length of stay (LOS) for ICU and non-ICU patients, as well as the expected percent of ICU patients on ventilators. The application then uses these inputs to calculate the fraction of newly admitted ICU, non-ICU, and ICU-ventilator patients expected each day.

Because bed needs must also reflect the numbers of beds needed for new admissions, those opened from discharges, and those currently occupied, our application models the daily carry-over of the patient census using expected LOS and the cumulative distribution function (cdf) of the binomial distribution.

The binomial distribution is a discrete probability distribution that models binary outcomes (e.g., patients either leave the hospital or stay for an additional day) and requires only two parameters (*p*, *n*). We set the value of *p* to 0.5 and set the value of *n* to be twice the average LOS. Doing so produces a symmetrical probability mass function (pmf), with a mean equal to the average LOS. This pmf is then converted to a cdf, which produces a first approximation for the fraction of 1-day, 2-day, …, etc., patients that are expected to be discharged on the present day. The fraction of patients not expected to be discharged are then carried over to the following day, e.g., a 1^st^-day patient becomes a 2^nd^-day patient and thereby has a different probability of leaving the hospital on the current day. This process is then iterated from the date of the first COVID-19 admit to the latest day in the user-requested forecast window (Fig 5).

**Figure 5.**
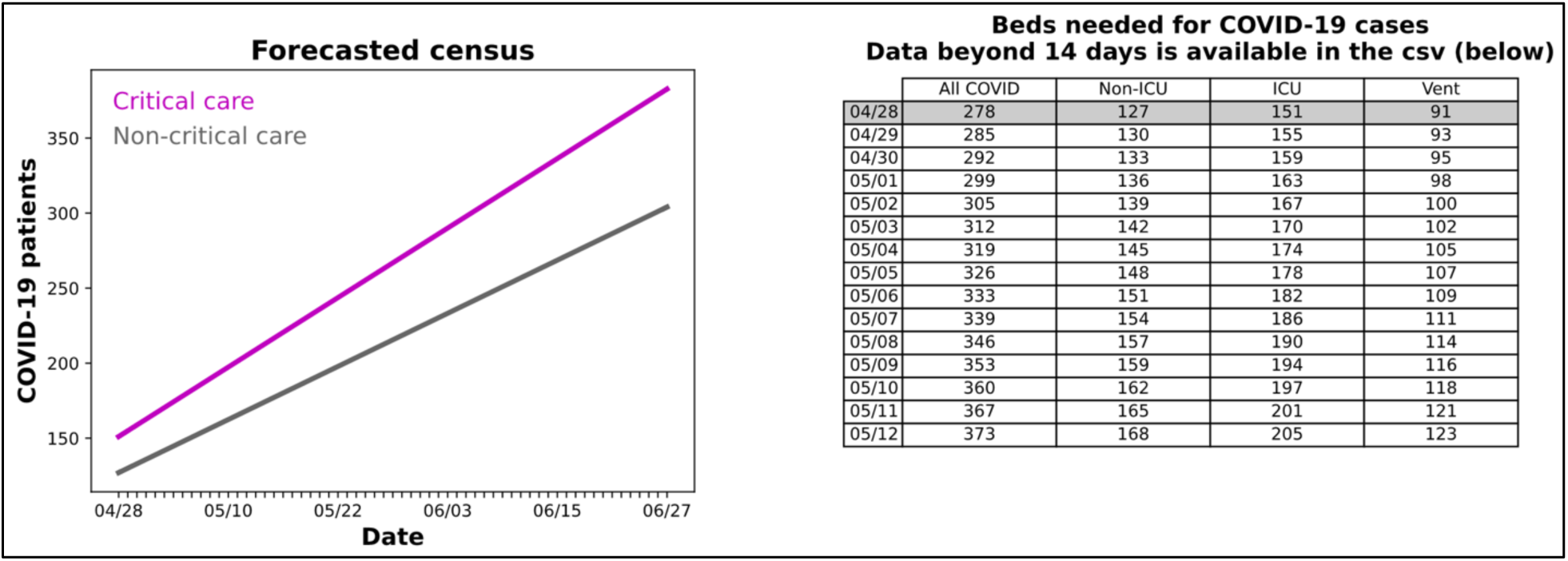
The third pane our application plots and tabulates the forecasted patient census. Left: A graph of forecasted numbers of critical care and non-critical care patients. Right: A table of forecasted bed needs for all COVID-19 patients, non-ICU patients, ICU patients, and ICU patients on ventilators. The application informs the user that forecasted data beyond 14 days are available via csv downloads.

### Forecasting supply needs

Our application allows users to forecast personal protective equipment (PPE) needs in accordance with the forecasted census of COVID-19 ICU, non-ICU, and ICU on ventilator patients. Users can enter expected per patient per day values for surgical gloves, nitrile exam gloves, vinyl exam gloves, anti-fog procedural face masks, fluid resistant procedural face masks, extra-large yellow isolation gowns, anti-fog (with film) surgical face masks, anti-fog full face shields, and particulate filter respirators. These values entered are then multiplied by the respective patient type across the forecasted patient census to produce graphical and tabulated results (Fig 6).

**Figure 6.**
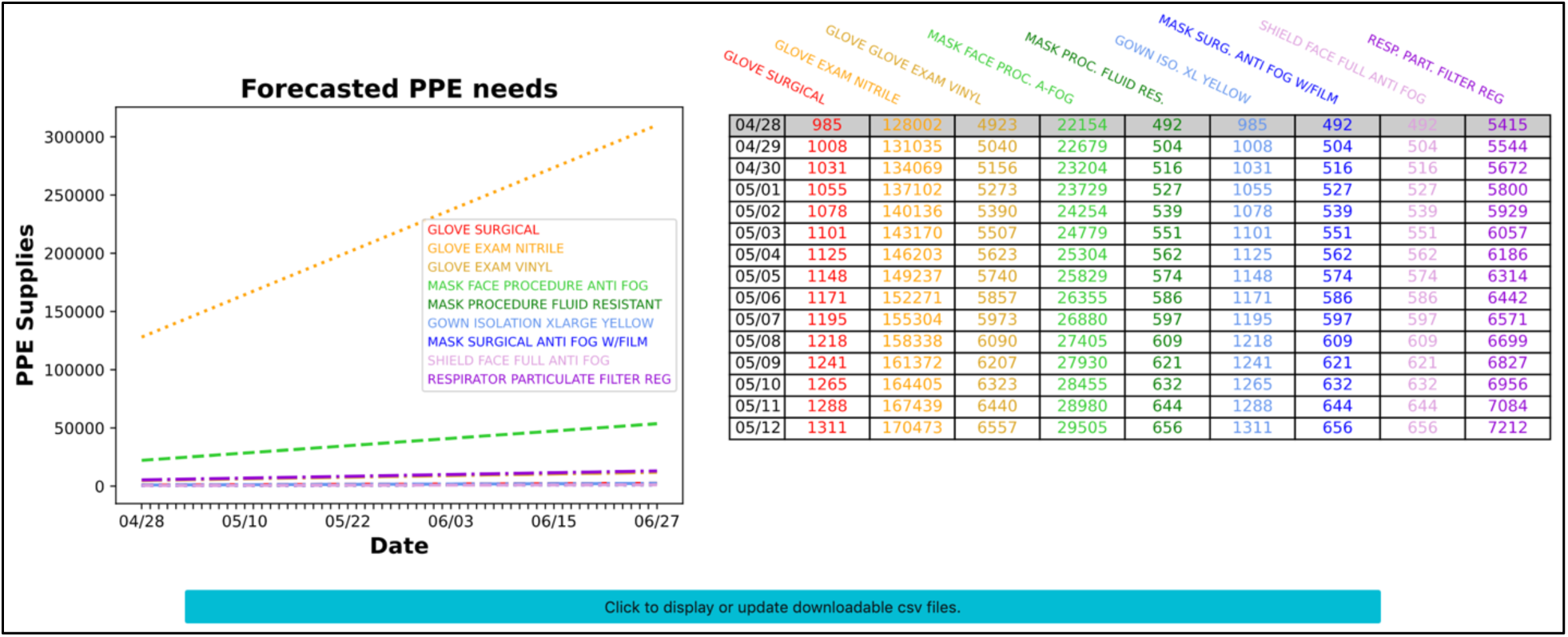
The bottom pane of our application plots and tabulates forecasted PPE needs. Left: Forecasted PPE needs are plotted to reveal overall expected trends. PPE supplies with similar input values become plotted on top of each other. Right: Forecasted PPE are tabulated with color coding that corresponds to the figure on the left, to allow easier visual interpretation. The application informs the user that forecasted data beyond 14 days are available via csv downloads. Users can click the teal button at the bottom to display or refresh downloadable csv files.

### Provisioning of forecasted data

Our application allows users to download forecast data in .csv format from each displayed table. This includes a table of forecasted cases, visits, and admits, a table of ICU and non-ICU bed needs, and a table of PPE supply needs. These downloadable files correspond to the model chosen by the user. Importantly, for layout purposes, the application does not display rows of data beyond a 14-day forecast. Instead, the user can view data from longer forecasts via the downloadable .csv files.

### Provisioning of source code

Source code for our application was built using the Python language (v3.7.4), Jupyter notebook, and the python-based Voila software which converts Jupyter notebooks to dashboard-like applications. All code and associated data are available from the public GitHub SupplyDemand repository, found on the Rush Quality Safety and Value analytics GitHub organization (https://github.com/Rush-Quality-Analytics/SupplyDemand). The repository provides an informative README.md file and the source code includes extensive commenting to assist users in their use and potential modification.

## DISCUSSION

### Meeting immediate needs

Hospitals and healthcare enterprises are endeavoring to make appropriate preparations and acquire adequate supplies to meet the challenges of the COVID-19 pandemic. While many studies have aimed to characterize the basic epidemiology of the disease, and many online tools have been developed to visualize its spread, few tools have been developed that empower hospitals to make informed decisions about expected visits and admits, ICU beds, ventilator, and PPE needs. To this end, our application is already in use by our home institution and several hospitals across the country and is available on the http://covid19forecast.rush.edu/ website. Consequently, the present work is intended to 1) make our application broadly known to the healthcare and scientific communities 2) give healthcare providers an in-depth understanding of our application, and 3) to point specialists, non-specialists, and in-house predictive analytics teams to a freely available tool that can also serve as modifiable platform of analytical source code and aggregated data. While some regions in the United States have started to experience peaks in cases with plateaus and declines, we anticipate that until a vaccine is available, episodes of repeated increased cases will be seen, for which forecast models will inform operational responses.

### Novel insights for applied and basic research

While our intention was not to provide *de novo* models or to provide refined epidemiological parameter estimates, our tool does allow for novel insights. First, our SEIR-SD model incorporates two phenomena of global, national, and local concern, i.e., social distancing and lags in COVID-19 testing. To our knowledge few extensions of the SEIR model have accounted for social distancing in being driven by an emergent social response to increased percent infected and as driven by external forces (e.g., public policy).

Likewise, few if any SEIR models have accounted for the influence of lags in testing on the apparent size of the infected population. Second, we envision that applied and basic research studies can be conducted using the downloadable data of our application and the freely available source code. Specifically, users can investigate any number of simple-to-complex relationships using the downloadable forecast data that results from our suite of models and which is offered alongside adjustable forecast windows, time-lags, lengths of stay, as well as other customizable parameters and aggregated data (population size, date of first reported infection, numbers of confirmed COVID-19 cases).

### Pending and potential modifications

We are continually improving the functionality and performance of our application to meet the predictive analytic needs of our home institution and broader healthcare community. In the near future, we will include models to predict the eventual decline of the pandemic and potential resurgence as social distancing guidelines and other mitigating policies are relaxed. We also look to provide the functionality to 1) examine regions outside the US, 2) examine county-level regions within the US, 3) include a greater array of supply needs and forecasts for numbers of providers and staff needed, and 4) allow providers to begin planning how and when to increase the number of elective surgeries and ambulatory visits. In building our open-source application from a small set of freely available and highly popular software tools, we expect that other researchers and healthcare analytics teams could readily pursue these and other improvements.

### Caveats and limitations

Our web application is versatile and easy to use. However, users should consider the following caveats and limitations of modeling. First, our application draws from a widely used COVID-19 dataset that may not reflect the true prevalence of COVID-19 within each US state and territory. While our application allows users to enter several parameters (e.g., % of infected visiting one’s hospital), these may change over time. Additionally, length of stay for ICU and non-ICU patients likely fluctuates and may not necessarily be binomially distributed. Similarly, time lags in hospital visits may not necessarily be Poisson distributed and may also fluctuate across time. PPE usage rates may also fluctuate based on changing hospital policies and supply shortages. Finally, our application does not account for differences in susceptibility to COVID-19 with respect to age nor the influence of comorbid conditions and sociodemographic factors.

## Data Availability

All code and associated data are available from the public GitHub SupplyDemand repository, found on the Rush Quality Safety and Value analytics GitHub organization (https://github.com/Rush-Quality-Analytics/SupplyDemand).

## ACKNOWLEDGMENTS

We thank the Johns Hopkins University Center for Systems Science and Engineering (JH CSSE) for continuing to provide daily updated data on the COVID-19 pandemic.

